# NanoRanger enables rapid single base-pair resolution of genomic disorders

**DOI:** 10.1101/2023.10.14.23296971

**Authors:** Yingzi Zhang, Chongwei Bi, Seba Saleh Nadeef, Sateesh Maddirevula, Mashael Alqahtani, Fowzan S. Alkuraya, Mo Li

## Abstract

Rare diseases affect around 350 million individuals globally, yet at least half of those with suspected Mendelian disorders remain without a precise molecular diagnosis despite advanced genetic testing using short read sequencing (SRS). Long-read sequencing (LRS) holds a promise in addressing this diagnostic gap although its clinical application is hampered by its complicated workflow, demanding sample requirements, and exorbitant cost. Genomic disorders represent an opportunity to demonstrate the unique capability of LRS. This study addresses the challenges of identifying missing disease-causing breakpoints in complex genomic disorders by employing multiple LRS strategies, including a novel strategy named <u>nano</u>pore-based rapid <u>a</u>cquisition of <u>n</u>eighboring <u>ge</u>nomic regions (NanoRanger). NanoRanger requires neither detailed prior genetic mapping nor large amounts of ultrahigh-molecular-weight DNA, and it stands out for its ease of use and ultra-rapid acquisition of large genomic regions of interest with deep coverage. We describe a cohort of 13 families, each harboring a different homozygous disease-causing genomic rearrangement that defied breakpoint determination by SRS and Optical Genome Mapping (OGM). In each case, NanoRanger identified the breakpoints with a single base-pair resolution. This has enabled the accurate determination of the carrier status of unaffected family members as well as the founder nature of these genomic lesions and their frequency in the local population. It has also enabled an unprecedented analysis of the DNA motifs to discern the mechanism that predisposed to these recessive rearrangements. Our data suggest that NanoRanger can greatly accelerate the clinical adoption of LRS and expand its access for the benefit of patients with rare diseases.

## MAIN

More than 350 million people worldwide have a rare disease^1^. 80% of rare diseases are estimated to have a genetic origin^1^. Despite advances in routine genetic testing in both clinical and research settings, approximately half of individuals worldwide suspected of having a Mendelian disorder remain undiagnosed^1–3^. For those fortunate enough to receive an accurate diagnosis, it takes an average of 4.8 years^1^. Up to 80% of rare disease patients will be misdiagnosed at least once ^4,5^. The low diagnostic yield and lengthy diagnostic odyssey take a heavy toll on the quality of life of the patients and their families.

The failure of exome sequencing (ES) to secure a molecular diagnosis in at least 50% of patients with suspected Mendelian diseases in large cohorts has been attributed by some to its limited coverage compared to genome sequencing. However, the literature suggests that the added value of genome sequencing over ES is minimal^6,7^. This suggests that disease-causing variants may have been equally detected by exome and genome sequencing but are missed at the interpretation level. Indeed, we have shown that interpretation accounts for a substantial fraction of “negative” cases^8,9^. The second possibility, not mutually exclusive with the first, is one of detection rather than interpretation i.e., the technology that exome and genome sequencing are currently based on (short read sequencing or SRS) is inherently limited in its analytical sensitivity. In other words, long read sequencing (LRS) may address the detection limitation of SRS. This may be particularly true in the case of genomic disorders, a term first coined by Lupski in 1998 to describe diseases that result from rearrangements of the human genome rather than from DNA sequence base changes^10^. There are indeed a few studies demonstrating the superiority of LRS over SRS in this class of disorders^11,12^. However, whole genome LRS is still prohibitively expensive for routine genetic diagnosis and remains largely inaccessible due to complex workflow and stringent sample requirement for ultrahigh molecular weight DNA. Optical Genome Mapping (OGM) has emerged recently as a potentially sensitive alternative for high resolution analysis of genomic disorders. However, cost and stringent sample requirements remain a major obstacle^8,13,14^.

While several methods including long-range PCR (LR-PCR), Targeted Locus Amplification (TLA), Cas9-mediated isolation of targets, and adaptive sampling, have been reported to enrich target genomic regions for LRS, none of them is generally applicable to different clinical scenarios due to different technical limitations ^11,15,16^. For instance, LR-PCR requires laborious primer design and optimization by trial and error due to amplicon size limit (<30kb) and uncertainty of the genetic lesion. TLA is limited by the requirements of millions of live cells and complex experimental processes. Computational T-LRS methods such as adaptive sampling necessitate a significant quantity of DNA material and sequencing resources to achieve adequate sequencing depth, rendering them impractically resource-intensive in its current format.

To overcome these limitations, we developed NanoRanger, which enables rapid and accurate detection of all types of SVs in expanded genomic regions at base resolution. Compared with other T-LRS methods, NanoRanger requires significantly less DNA and a fraction of the flow-cell capacity while achieving tens of thousands fold higher sequence coverage. We first evaluated current T-LRS approaches using four clinical cases with unresolved breakpoints and compared their performance with that of NanoRanger. We then used NanoRanger to resolve nine cases of suspected autosomal-recessive Mendelian disorders, which all failed to be diagnosed with conventional genetic tests (e.g., molecular karyotyping by chromosomal microarray, ES, and OGM). Using the validated breakpoints, we screened for carriers in 1000 healthy Saudi individuals and performed in-depth analysis of the DNA motifs at the breakpoints to explore the potential mechanism. Our results demonstrate that NanoRanger is a rapid and cost-effective approach with wide clinical applicability.

## RESULTS

To systematically explore the utility of T-LRS in genetic diagnosis we assembled a cohort of Saudi patients (Table 1) who had been diagnosed with diverse recessive genetic disorders but all lacked accurate molecular diagnoses. The study encompassed a wide spectrum of disease pathologies commonly found in large Mendelian genomics programs, including neurodevelopmental disorders, dysmorphic/congenital malformation syndromes, inborn errors of metabolism, hematological conditions, immunological disorders, ophthalmological diseases, audiological disorders, pulmonary conditions, gastrointestinal issues, connective tissue-related disorders, cardiovascular diseases, skeletal abnormalities, reproductive disorders, and renal conditions.

**Table 1.**
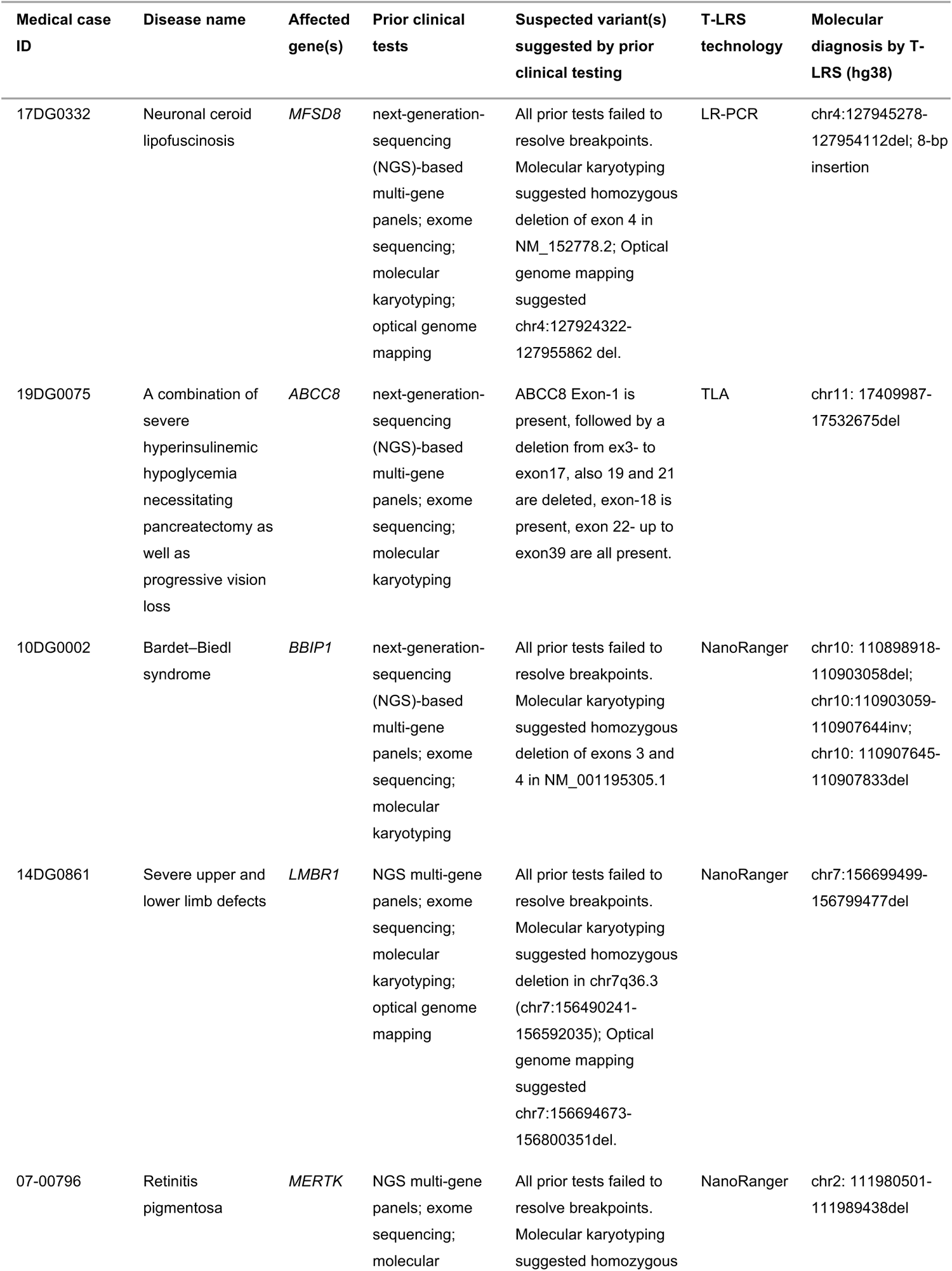
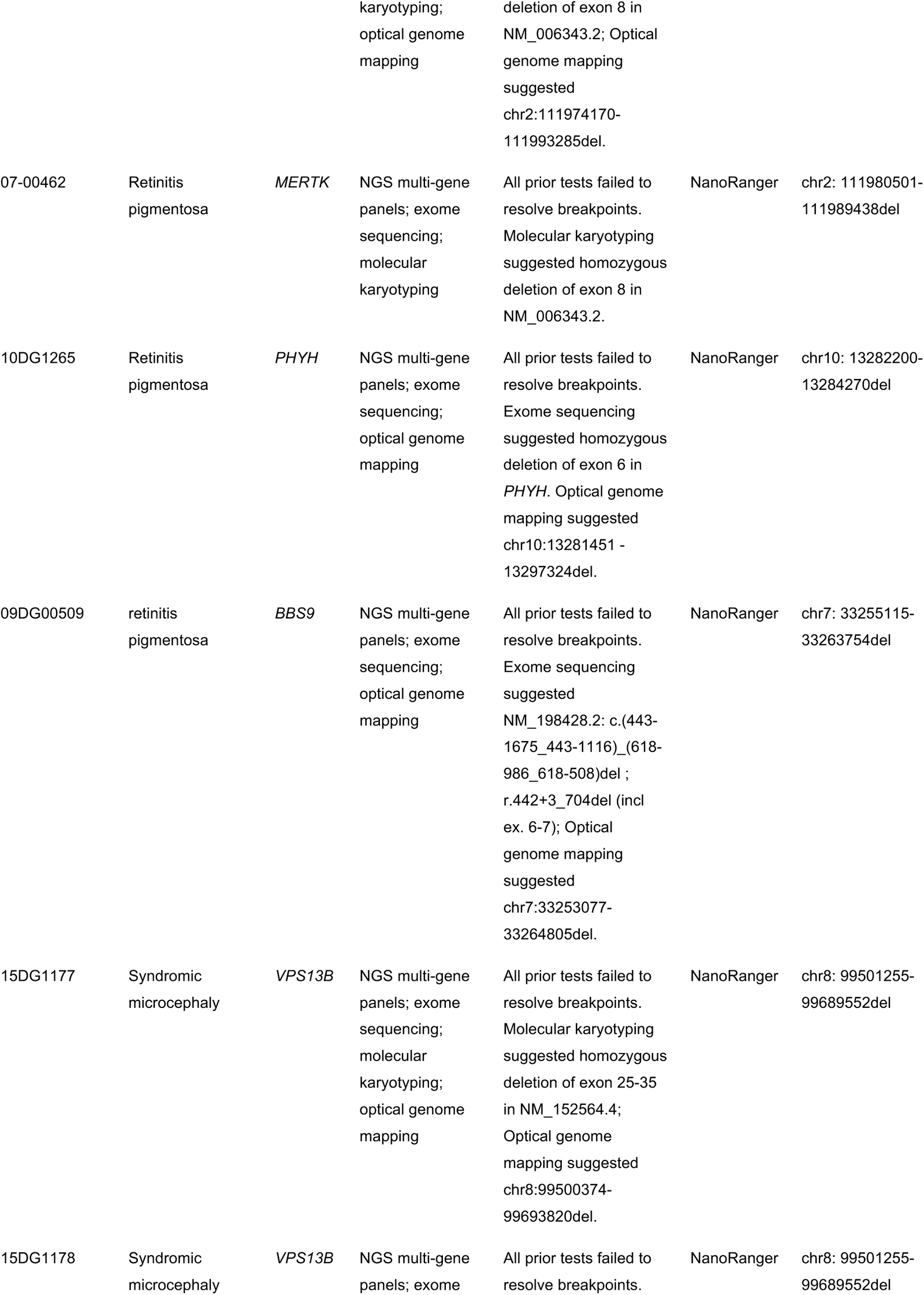
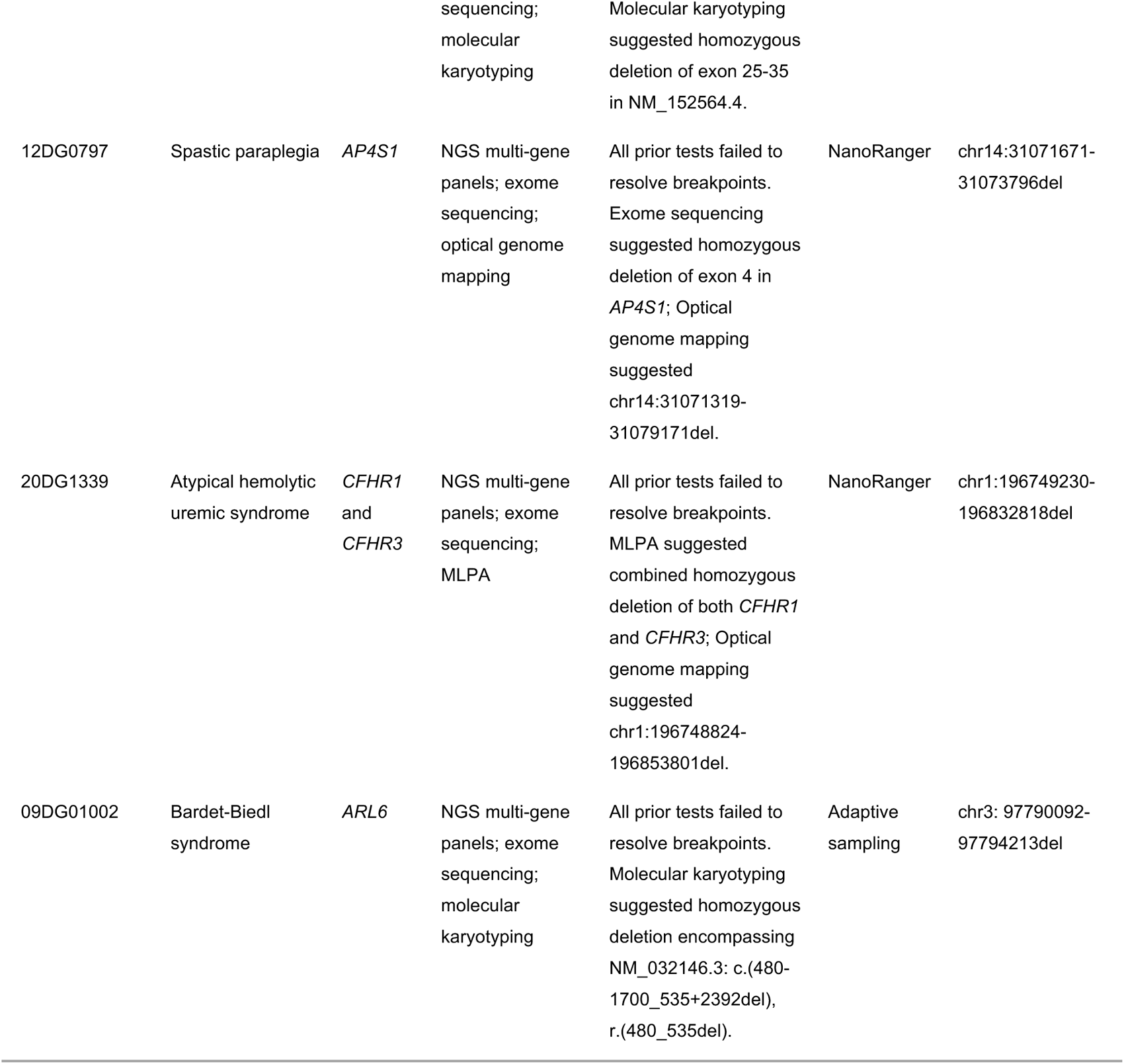
Case summary.

### LR-PCR and Nanopore sequencing identify large SVs that were missed by conventional clinical testing

In a male case of neuronal ceroid lipofuscinosis (17DG0332, Table 1), the initial diagnosis by molecular karyotyping indicated a homozygous deletion of exon 4 in the *MFSD8* gene, but the exact breakpoints could not be identified. To define the breakpoints, a long-range primer pair was designed for PCR amplification of the locus in question (refer to Supplementary Table 1 for details). The amplicon was pooled and sequenced on an Oxford Nanopore MinION flow cell. The 84,929 reads aligned to the MFSD8 gene in the hg38 revealed an 8.8-kb deletion (chr4: 127945278-127954112del) encompassing exon 4 (Fig. 1a). De novo assembly generated a 5006-bp consensus sequence that further revealed an 8-bp insertion at the deletion breakpoint, the exact sequence of which was verified by Sanger sequencing (Fig. 1b-c). The SV identified by T-LRS in *MFSD8* is novel and not currently listed in the ClinVar database.

**Fig. 1.**
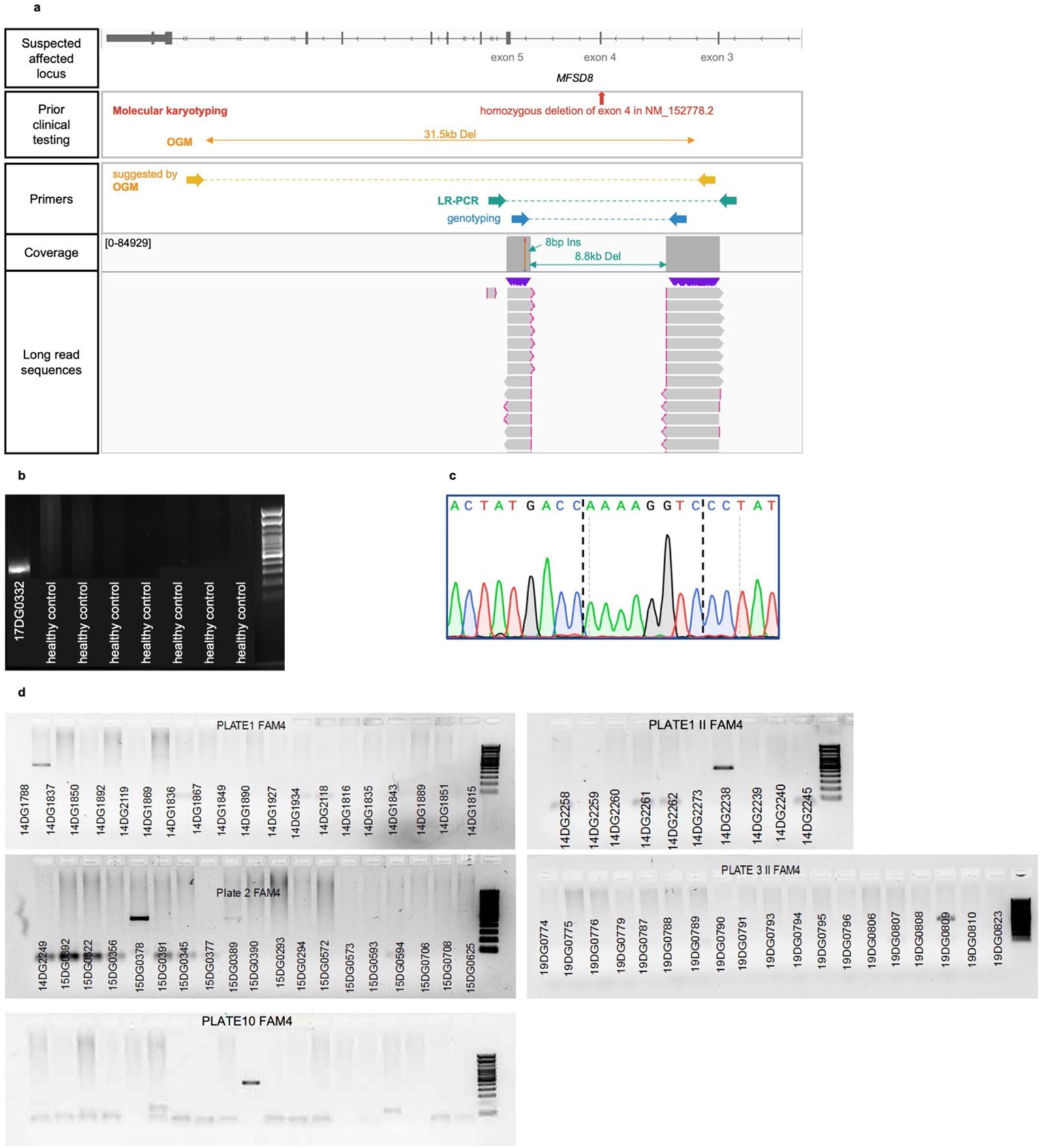
One medical case resolved by LR-PCR and Nanopore sequencing. **a,** A schematic summary of the gene map, prior test results, primer design, and sequencing results. **b,** Genotyping PCR identified the affected patient and the family members from healthy controls. **c,** Sanger sequencing validated the breakpoint identified by Long-range PCR and Nanopore sequencing. **d,** The genotyping primer pair was applied in a carrier screening of 1000 Saudi individuals. 5 out of 1000 individuals were revealed unaffected carriers of the MFSD8 breakpoint, suggesting a relatively elevated prevalence of the breakpoint within the healthy Saudi population. Only gels showing positive bands are presented.

OGM using Bionano technology was conducted in parallel. Bionano analysis suggested a 31.5 kb deletion (chr4:128845477-128877017 in hg19, transferred to chr4:127924322-127955862 in hg38) but could not provide the breakpoint sequence. Moreover, primers suggested by the Bionano breakpoints failed to produce any PCR product. After T-LRS resolved the SV, it became clear that Bionano analysis misplaced the proximal breakpoint by 1.8 kb and the distal breakpoint by 21.0 kb, which misled the primer design (Fig. 1a).

A carrier screening for the *MFSD8* SV was performed in 1,000 healthy Saudi individuals (genotyping PCR primer sequences listed in Supplementary Table 1). The screening identified five carriers of the SV, which establishes this as a previously unrecognized founder variant in the Saudi population (Fig. 1d).

### TLA and Nanopore sequencing identify large SVs missed by clinical testing

Despite the one success of the LR-PCR strategy, it failed to resolve most cases in our cohort. The failures lay in the difficulty in designing effective PCR primers to amplify the genomic region containing the unknown mutation. Typically, several pairs of primers are designed to flank the suspected lesion in increasing distances, but the range covered by this approach is limited by the upper limit of LR-PCR (∼30 kb, beyond which positive control cannot be obtained, making it impossible to interpret negative PCR results). This strategy therefore is ineffective for large SVs, as the trial-and-error approach would consume excessive labor, time, and samples. TLA, on the other hand, can capture a locus of up to a few hundred kilobases in one test, thus offering a more efficient strategy to resolve large SVs.

One case in our cohort has a potential founder deletion in Arab population, affecting portions of the *USH1C* and *ABCC8* genes. Patients with this homozygous deletion exhibited a complex clinical picture, combining severe hyperinsulinemic hypoglycemia—which necessitated pancreatectomy—with progressive vision loss. However, the absence of precise coordinates for this deletion hindered the development of a screening assay for unaffected carriers, perpetuating the risk of having children afflicted by this debilitating combination of symptoms. Molecular karyotyping of one male case (19DG0075, Table 1) initially suggested a homozygous deletion of exons 3-17 and 19-21 of *ABCC8*, while exons 1, 18, and 22-39 appeared to be present. Several LR-PCR attempts failed to amplify specific products from patient DNA. Fortunately, a lymphoblastoid cell line (LCL) was available for this case, which allowed TLA using primers anchored flanking the existing regions, positioned away from the suggested deletion, followed by Nanopore sequencing. Analysis of the nanopore reads captured by TLA showed a deep coverage spanning 318-kb around the *ABCC8* locus and an absence of reads in a 123-kb region encompassing parts of the *ABCC8* and *USH1C* genes, suggesting a large deletion (Fig. 2a). The breakpoint was verified by genotyping PCR and Sanger sequencing (Fig. 2b-c). The genotyping assay successfully identified the carrier siblings and was used to screen for carriers in 1000 healthy Saudi individuals (Fig. 2d).

**Fig. 2.**
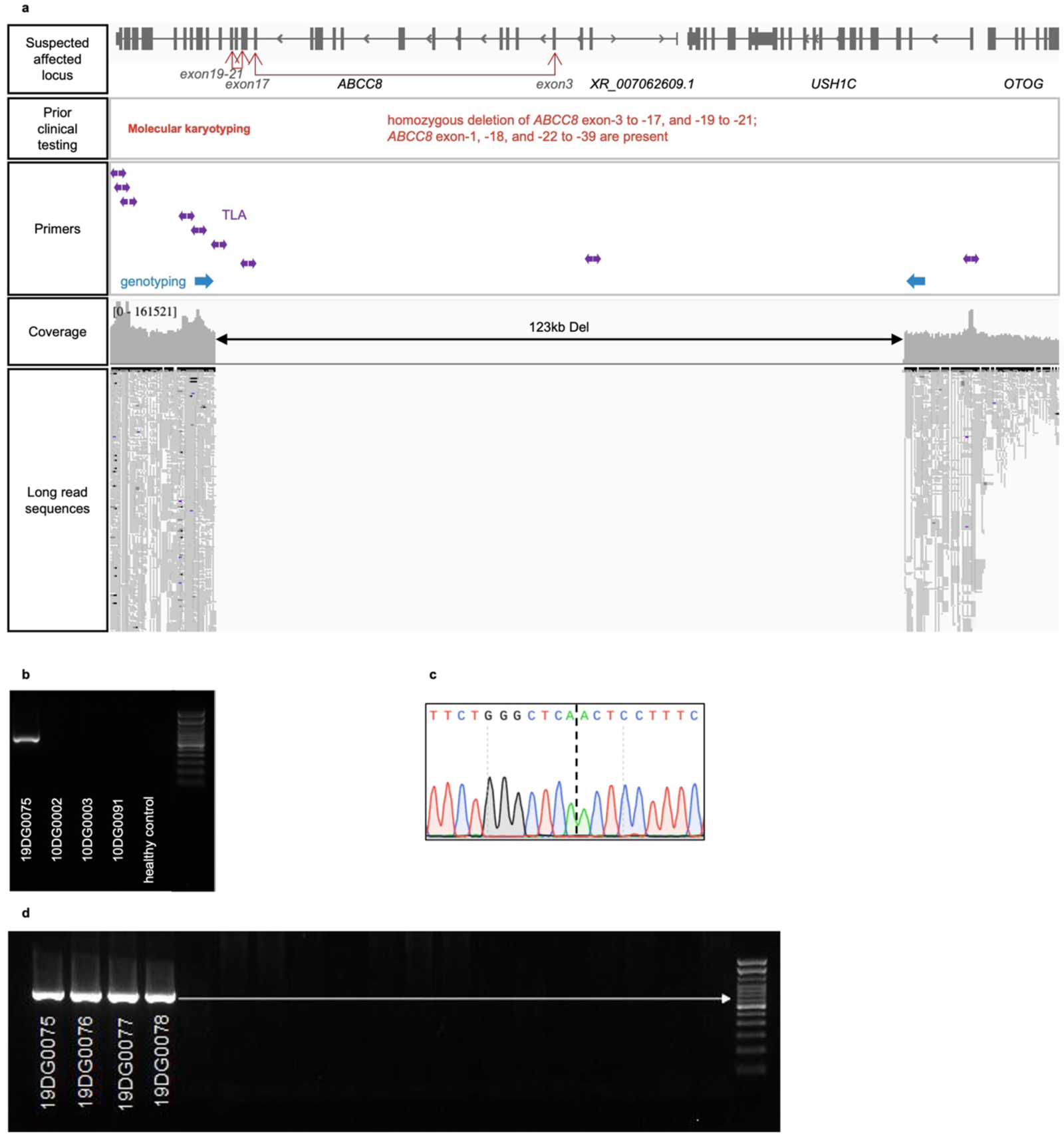
One medical case resolved by TLA and Nanopore sequencing. **a,** A schematic summary of the gene map, prior test results, primer design, and sequencing results. **b,** Genotyping PCR identified the affected patient from the unrelated families or healthy controls. **c,** Sanger sequencing validated the breakpoint identified by TLA and Nanopore sequencing. **d,** The genotyping primer pair was applied in a carrier screening of 1000 Saudi individuals. The carrier siblings were successfully identified. Only gels showing positive bands are presented.

This case study demonstrated the advantage of TLA over LR-PCR in efficiently capturing and amplifying large genomic regions (up to hundreds of kilobases), thereby facilitating the detection of large deletions. However, TLA as a genetic testing strategy has several intrinsic limitations. Firstly, the capture frequency (i.e., sequencing coverage) of any DNA fragment by the anchor depends on their physical proximity fixed by crosslinking, which means copy number variations and duplications will be difficult to interpret. Secondly, TLA reads are from ligation-mediated PCR of crosslinked restriction-digestion fragments and thus require split read mapping, which confounds analysis of many types of SVs such as inversions, translocations, and complex rearrangements. Lastly, TLA requires a large number (5-10 million) of live cells and a long and complex workflow that typically lasts 5-7 days–both unrealistic demands in clinical settings.

### NanoRanger accurately and rapidly identifies complex SVs

We reasoned that an ideal T-LRS strategy should retain the efficiency of TLA but avoid its pitfalls. Since most issues of TLA discussed in the preceding paragraph originate from the crosslinking of chromatinized DNA, we invented a strategy that rapidly captures large neighboring genomic regions without crosslinking. This strategy, when coupled with nanopore sequencing is called <u>nano</u>pore-based rapid <u>a</u>cquisition of <u>n</u>eighboring <u>ge</u>nomic regions (NanoRanger).

NanoRanger offers targeted sequencing of extensive genomic regions—ranging from tens to hundreds of kilobases—in the candidate locus. This is achieved through a combination of partial restriction digestion, ligation-mediated inverse PCR, and long-read sequencing (Fig. 3).

**Fig. 3.**
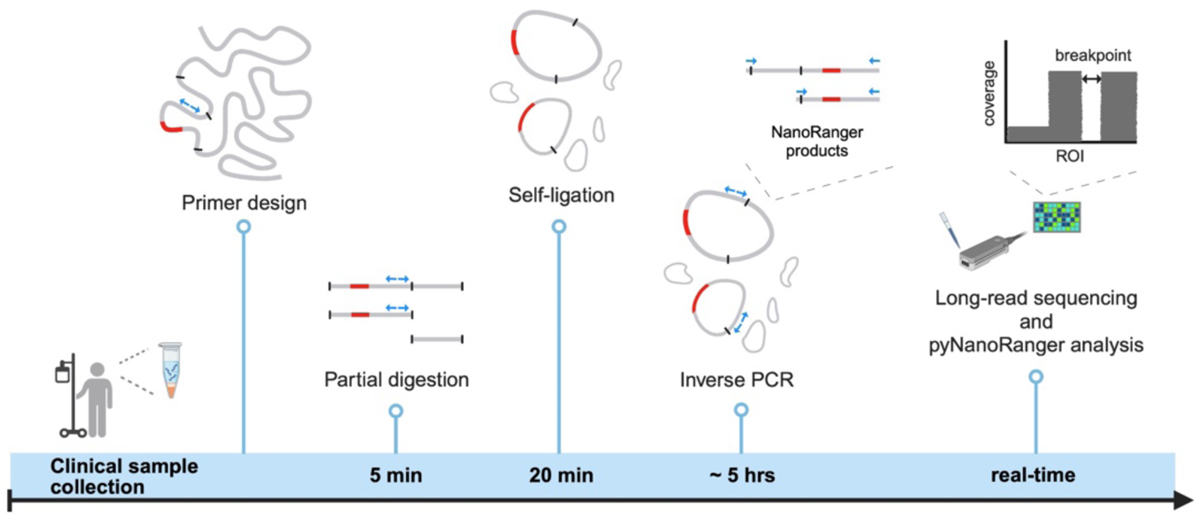
Schematic representation of NanoRanger. DNA is extracted from the collected clinical samples. Individual DNA molecules containing the genomic region of interest (ROI, red color) are digested, self-ligated, and amplified for sequencing on appropriate platforms (e.g., Illumina, PacBio and Nanopore). The sequencing data are analyzed by a bioinformatic toolkit-pyNanoRanger-to identify amplicon sequences, bin reads based on digestion sites and call variants. Thick gray lines: genomic DNA. Blue arrows: inverse PCR primers. Black bars: restriction enzyme sites.

First, inverse PCR primers are designed within expected wild-type sequences (the “anchor” sequences) in the sample (through a priori knowledge or testing) that are near the suspected mutation locus. The design is highly flexible so the tentative breakpoints can be of various distances from the primers. Genomic DNA is extracted from patient samples using routine methods *without* crosslinking. An appropriate restriction enzyme is then selected to digest the genomic DNA into fragments typically ranging from a few kilobases to tens of kilobases. The enzyme is allowed to work briefly (∼ 5 min) so that partial digestions allow for the production of restriction fragments of diverse sizes. The resulting restriction fragments are self-circularized using DNA ligase under conditions strongly favoring intra-molecular ligation^17^. These circularized restriction fragments are then used for inverse PCR amplification of DNA regions adjacent to the suspected breakpoints. Circularized restriction fragments of different sizes (from the deliberate partial digestions) are amplified by the same primers to improve the coverage of the locus. This strategy eliminates the tedious trial-and-error primer testing in LR-PCR, because any changes to the wild-type restriction sites, be it elimination due to deletions or creation due to rearrangements, would be captured by the partial digestion, self-circularization, and inverse PCR steps. Finally, the amplicons are sequenced using long-read sequencing technologies (Fig. 3). A bioinformatics pipeline called pyNanoRanger is tailored to handle NanoRanger sequencing data analysis (see Methods) (Fig. S1).

We tested the efficacy of NanoRanger using a male case of Bardet–Biedl syndrome (10DG0002, Table 1). Previous testing by molecular karyotyping suggested a homozygous deletion of exon 3 and 4 (based on accession number NM_001195305.1) without definitive breakpoint coordinates. Inverse PCR primers were designed for the suspected mutation locus (Supplementary Table 1). We selected PstI as the restriction endonuclease for partial digestion. Two hundred nanograms of genomic DNA samples of the proband and his parents (collectively termed the “trio” in this context) were processed using the NanoRanger protocol (see Fig. 3). Gel electrophoresis of the inverse PCR products revealed a different pattern of bands in the trio samples as compared with a healthy control (Fig. 4b). The PCR products were purified and sequenced on a MinION flow cell. The sequencing data were processed and analyzed using pyNanoRanger.

**Fig. 4.**
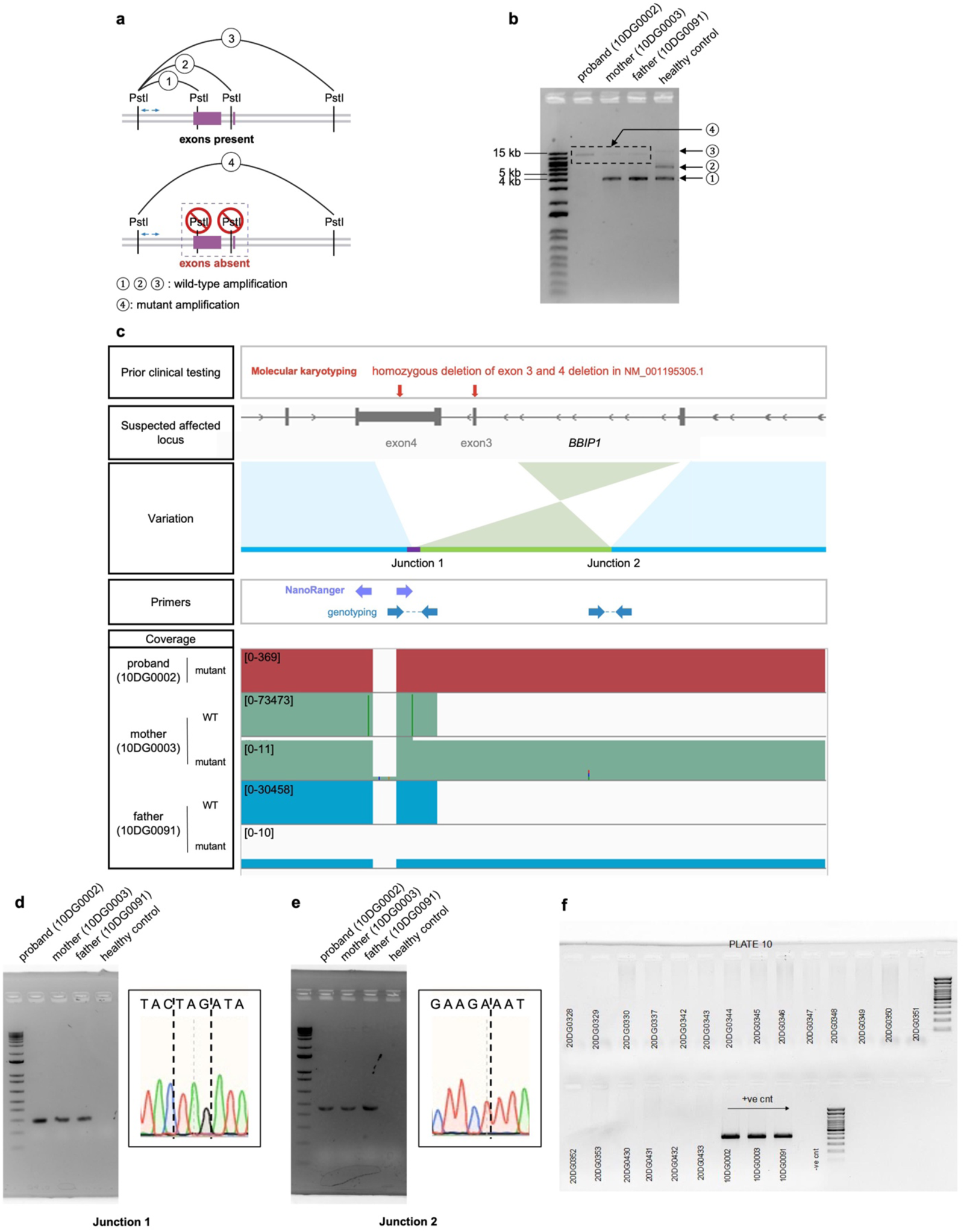
NanoRanger enables precise characterization of disease-causing breakpoints in a Bardet-Biedl Syndrome case (10DG0002). **a**, Schematic illustration of the NanoRanger strategy for the region of interest. **b,** Gel electrophoresis results of Inverse PCR products, displaying the mutant band in affected family samples and wild-type bands in the parents and a healthy control. **c,** A schematic summary of the gene map, prior test results, primer design, and sequencing results. **d, e,** The breakpoint junctions 1 (d) and 2 (e) were verified by Sanger sequencing. **f,** The genotyping primer pair for Junction 1 was applied in a carrier screening of 1000 Saudi individuals. The carrier parents were successfully identified. Only gels showing positive bands are presented.

The long reads facilitated sequence alignment and phasing, which revealed that the proband is homozygous for the mutant haplotype, whereas the parents each has one mutant allele and one wild-type allele (Fig. 4c). De novo assembly using NanoRanger generated a 12-kb consensus sequence, which shows a complex rearrangement comprising a 4.6-kb inversion, a 189-bp deletion, and a 4.1-kb deletion (Fig. 4c). The breakpoint junctions were further verified by Sanger sequencing (Fig. 4d). NanoRanger provided high sequencing coverage that precisely pinpointed the breakpoints at base-level resolution. This proof-of-concept experiment demonstrated that NanoRanger can efficiently target DNA regions suspected of containing unmapped inherited disease variants, facilitating the resolution of large SVs in patients with genetic diseases.

A genotyping assay based on the NanoRanger breakpoints successfully confirmed the carrier status of the parents. We further designed a multiplex genotyping assay based on the resolved breakpoints of *MFSD8*, *ABCC8* and *BBIP1* and conducted the genotyping assay in 1,000 healthy individuals. The assay successfully identified the carrier siblings of each breakpoint (Fig. S6). The results demonstrate the effectiveness of multiplex genotyping for at least three pathogenic breakpoints, highlighting the value of this study to provide cost-effective premarital carrier screening for novel genetic disorders.

### NanoRanger is widely applicable to diverse clinical cases

Next, we asked if the success of NanoRanger is generalizable to different gene loci and to SVs of various sizes. To this end, we focused on the remaining cases from the Saudi genetic disorder cohort (Table 1). These individuals had been diagnosed with a range of recessive genetic disorders but lacked precise molecular diagnoses because the coordinates of their rearrangements could not be determined by other technologies.

An individual (09DG00509) with Barde-Biedl syndrome had previously undergone a series of diagnostic tests, including NGS multi-gene panels, ES, and OGM. Unfortunately, none of these tests succeeded to pinpoint the breakpoints. ES suggested a deletion encompassing exons 6 and 7, while OGM indicated a deletion approximately 11kb in length) (Table 1, Fig. S2). With NanoRanger we identified a 8,640-bp deletion involving the *BBS9* gene (Table 1, Fig. S2). Remarkably, it took only 40 minutes of nanopore sequencing to uncover the breakpoint with a read depth of 10,973, showcasing the efficiency of NanoRanger in resolving complex cases.

In the female case labeled 14DG0861, characterized by severe developmental defects in both upper and lower limbs, a series of diagnostic tests including NGS multi-gene panels, ES, molecular karyotyping, and OGM were unsuccessful in identifying the breakpoints. Molecular karyotyping of the affected individual indicated a homozygous deletion in chr7q36.3, approximately 100kb in length, encompassing *LMBR1* (NM_022458.3), while OGM suggested a deletion 106 kb in length (Table 1). Genotyping PCR based on these suggested breakpoints failed to produce any product. NanoRanger identified a 99,979-bp deletion at chr7:156699499-156799477, which was verified by Sanger sequencing (Fig. S3).

Similarly, NanoRanger identified base-resolution breakpoints in seven more cases that had undergone various conventional genetic tests (e.g., NGS multi-gene panels, ES, molecular karyotyping, OGM, and Multiplex Ligation-dependent Probe Amplification (MLPA)) but remained unresolved. These included two male cases (07-00462 and 07-00796) of retinitis pigmentosa with a 8,938-bp deletion involving the *MERTK* gene, one male case (10DG1265) of retinitis pigmentosa with a 2,071-bp deletion involving the *PHYH* gene, two male cases (15DG1177 and 15DG1178) of syndromic microcephaly (Cohen syndrome) with a 188,298-bp deletion involving the *VPS13B* gene, one male case (12DG0797) of spastic paraplegia with a 2,126-bp deletion involving the *AP4S1* gene, and one male case (20DG1339) of atypical hemolytic uremic syndrome with a 83,589-bp deletion involving the *CFHR1* and *CFHR3* genes (Table 1)^18^.

Together, these cases underscore the universal applicability of NanoRanger in elucidating complex genetic anomalies that eluded conventional genetic testing.

### Adaptive sampling vs. multiplex NanoRanger

Oxford Nanopore sequencing offers a unique method called adaptive sampling that takes advantage of the independent voltage control of individual sequencing channels and real-time software analysis of sequencing results to achieve target enrichment in native DNA^11,19^. Because the targets can be added in the software with virtually no additional cost, adaptive sampling has the potential to be a simple and efficient T-LRS solution. To compare adaptive sampling with NanoRanger in a realistic clinical scenario, we applied adaptive sampling to the same Bardet–Biedl syndrome case (10DG0002) resolved by NanoRanger. To simulate de novo diagnosis, we chose 16 Bardet-Biedl gene loci as enrichment targets. (Supplementary Table 2). In a preliminary experiment in 293T cells, Average coverage of the 16 target regions ranged from 10.0-44.9x. The average read length in the interested gene regions ranged from 5,933 bp to 13,614 bp (Supplementary Table 2). We then applied adaptive sampling to 10DG0002 sample. Average coverage of the 16 target regions ranged from 2.3x to 8.4x. The average read length ranged from 1854 bp to 2913 bp (Supplementary Table 3). Despite the low sequencing depth and shorter reads, adaptive sampling reads showed the correct breakpoints as identified by NanoRanger (Supplementary Table 3) (Fig. S4a). However, the two junctions were covered by only three and four reads, respectively, making it difficult to confidently call the true sequence of the SV solely from adaptive sampling (Fig. S4b). Another Bardet–Biedl syndrome case (09DG0002, Table 1) was similarly tested. Average coverage of the 16 target regions ranged from 7.3x to 11.1x; the average read length ranged from 1450 bp to 2035 bp (Supplementary Table 4). Adaptive sampling revealed a large deletion in *ARL6* covered by four reads, later validated by Sanger sequencing (Table 1) (Fig. S5a-b). The adaptive sampling process consumed ∼1 μg (vs. 200 ng for NanoRanger) of DNA, up to two (vs. 10% of the capacity of one MinION flow cell for NanoRanger) MinION flow cells, and up to 48 hours (vs. 40 mins for NanoRanger) of sequencing, which proved too resource intensive to be practical in its current format.

To extend the successful performance of NanoRanger in single locus to multiple loci, we tested multiplexed NanoRanger in one reaction. We chose the inverse primers for the *ABCC8* and *BBIP1* loci, known for their efficacy in TLA and NanoRanger, respectively (Supplementary Table 1). We utilized DNA samples extracted from wild-type 293T cells and tested four primer concentrations (Supplementary Table 5). The gel electrophoresis of the resulting amplicon products revealed consistent patterns. The PCR products were then purified, barcoded, and sequenced on a MinION flow cell. While total read count generally increased with primer concentration, the effective sequencing depth peaked at the 0.2 uM primer concentration (Supplementary Table 5). Under this condition, NanoRanger covered a span of 10 kb in *BBIP1* region with 7448 reads and 15 kb in *ABCC8* region with 3379 reads. The results demonstrate the effectiveness of multiplex NanoRanger for at least two gene loci, highlighting its potential as a first-line test for genetic disorders with candidate genes and as a cost-effective premarital carrier screening test for genetic disorders.

### Molecular mechanisms of novel recessive breakpoints

In our comprehensive analysis of breakpoints across the spectrum of diseases, we observed a predominant association with mechanisms influenced by repetitive elements or those involving Microhomology-Mediated End Joining (MMEJ) (Table 2). In the case of neuronal ceroid lipofuscinosis (17DG0332), we found two *Alu* elements with identical orientation near the 5’ and 3’ breakpoints, respectively, of the deleted DNA in the reference genome. This observation suggests that the deletion is mediated by nonallelic homologous recombination (NAHR) between the two homologous *Alu* elements, which is reminiscent to a previous report^20^. Additionally, for all four cases of retinitis pigmentosa, although the breakpoints are localized in different genomic regions, microhomology is consistently identified at the exact edges of all breakpoints. This observation suggests that the breakpoints were processed by microhomology-mediated end joining (MMEJ), highlighting the role of microhomology in shaping the genetic structural variations in retinitis pigmentosa^21^. T-LRS strategies–NanoRanger especially, have demonstrated remarkable efficacy in detecting and characterizing these elusive breakpoints, providing valuable insights into the upstream molecular pathways implicated in the pathogenesis of these disorders. The findings shed light on the underlying molecular processes contributing to the formation of these breakpoints^22^.

**Table 2.**
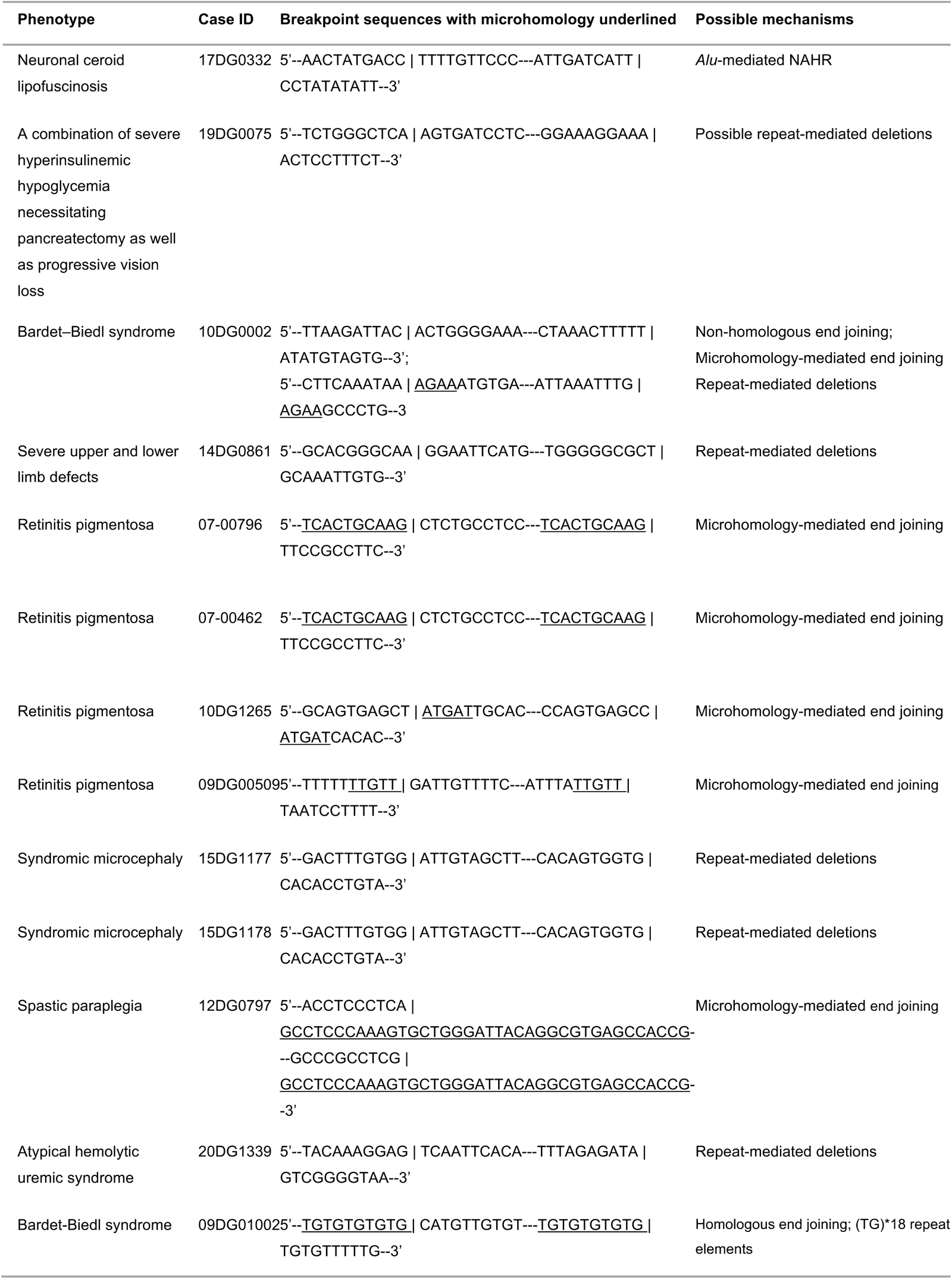
Breakpoints and the possible mechanisms.

## DISCUSSION

One of the vexing challenges in rare diseases is that we are only halfway towards the declared goal of granting every patient with a rare disease their right to an accurate molecular diagnosis within a year of presentation^23,24^. The fact that the diagnostic odyssey continues unabated even after whole-genome sequencing suggests that new approaches need to be implemented and the use of LRS represents one such approach. In this study, we aimed to address the challenges encountered by conventional approaches in genetic disease diagnosis. Our strategies take advantage of targeted sequence enrichment and LRS. Unlike many commercial SRS-based solutions, our strategies cover extensive spans of genomic regions of interest with long reads and deep coverage. We developed a simple and effective approach, NanoRanger, which shows paradigm-shifting reliability in resolving disease-causing breakpoints that are missed by conventional clinical testing. NanoRanger works with routine clinical samples (e.g., peripheral blood) and resolves large or complex genomic alterations at base-pair resolution.

While the breakpoints we unveiled in our clinical studies are individually rare in the general population, their clinical identification holds significance for unaffected carrier diagnoses within the population and contributes to ongoing scientific endeavors related to the genes involved. It is worth noting that our study included DNA samples dating back up to 16 years. Some samples are of low quantity and/or quality that make them unsuitable for most LRS methods. Despite this suboptimal quality of the starting material, NanoRanger exhibited robust performance in detecting breakpoints and providing deep coverage in expanded regions (up to 22 kb in the resolved cases in this study). This underscores the competitive edge of NanoRanger over other technologies (Table 3). LR-PCR necessitates detailed prior locus information. While it can achieve high sequence coverage, it can only cover a limited range by trial and error and has a low success rate. TLA covers large spans with good depth. However, its crosslinking strategy confounds analysis of many types of SVs such as inversions, translocations, and complex rearrangements. The necessity for millions of live cells and a long and complex workflow makes it impractical in routine clinical testing. Adaptive Sampling is noteworthy for its flexibility of genomic coverage. However, it requires over 1 ug of DNA and a significant amount of sequencing time and resources, yet provides only shallow coverage.

**Table 3.** Comparison among T-LRS methods.

|  | **NanoRanger** | **LR-PCR** | **TLA** | **Adaptive**<br>**sampling** |
| --- | --- | --- | --- | --- |
| Knowledge from<br>prior testing | optional | required | required | optional |
| Sample<br>requirement | ~200 ng DNA | ~50 ng DNA | 5-10 million live<br>cells | ≥1 ug DNA |
| PCR | yes | yes | yes | no |
| Restriction<br>digestion | partial (~5 min) | no | complete<br>(overnight) | no |
| Success rate | high | low | intermediate | high |
| Range | hundreds of kb | < 30 kb | a few hundred kb | hundreds of kb |
| Sequencing<br>depth | high | high | high | very low |
| Time | ~ 6 hours | hours to days | 5-7days | 2~3 days |
| Cost | low | low | intermediate | very high |

NanoRanger in its current form has several limitations. Firstly, its range is limited by the maximal DNA lengths that can be amplified using PCR. This could pose challenges in resolving very large structural variations (e.g., extensive gene duplications) and may necessitate multiple rounds of NanoRanger. Secondly, PCR amplification erases DNA modifications, limiting its utility for simultaneous analysis of SVs and DNA methylation, as required especially in diagnosis scenarios for imprinting diseases. We are currently developing NanoRanger to enable detection of DNA modifications. Lastly, NanoRanger by design is limited to targeted rather than untargeted diagnostic scenarios. However, future iterations may overcome this limitation especially given the compatibility of NanoRanger to multiplexing. We have showed that NanoRanger for different gene loci could be multiplexed in one reaction. In further development, higher levels of multiplexing could be used to build gene panels (e.g., genetic disease and cancer genes). We anticipate that multiplex NanoRanger could serve as a first-line diagnostic tool for genetic disorders with well-defined candidate genes and as a cost-effective premarital screening test for common genetic disorders.

## MATERIAL AND METHODS

### Human subjects

All human subject samples were collected from King Faisal Specialist Hospital & Research Center (KFSHRC). The research complies with all relevant ethical regulations at KFSHRC. The study of human samples collection was approved by the Institutional Review Board (IRB) of KFSHRC and KAUST Institutional Biosafety and Bioethics Committee (IBEC). Patients with a suspected Mendelian disease are included in the Mendelian genomics cohort. Informed consent is obtained prior to enrollment. The consent covers the generation and use of patient-derived cell-lines (LCLs and Fibroblasts) whenever it is needed. The case IDs used in the study are research IDs that are not known to anyone outside of the research group.

### Prior testing strategies

Cases recruited prior to 2012 were analyzed using next-generation-based multi-gene panels relevant to their clinical phenotype. Negative cases and cases recruited after 2012 were submitted for ES. Select negative cases after ES were analyzed using optical genome mapping. MLPA was requested clinically as appropriate by the ordering physicians from various CAP-accredited laboratories, and their results were recorded. Supplemental Methods explain the technical details of the testing platforms.

### DNA isolation

DNA applied for Nanopore sequencing was isolated from fibroblast cells using standard methods. Fixed lymphocyte cell lines were prepared and preserved for TLA-based Nanopore sequencing.

### Long-range PCR

Each PCR reaction solution (50 µl) consisted of ∼50 ng genomic DNA, 10 μl 5X PrimeSTAR GXL Buffer, 4 μl dNTP Mixture, 1 µl primer each (final concentration 10pmol each) (see Supplementary Table 2), 1 μl PrimeSTAR® GXL DNA Polymerase, and sterile purified water to 50 μl. The PCR parameters were set following the manufacturer’s instructions. PCR products were qualified with gel electrophoresis and purified by AMPure XP beads (Beckman Coulter, cat. no. A63882).

### TLA

TLA was performed as previously described^15^. Briefly, patient derived cells were crosslinked by formaldehyde followed by restriction enzyme NlaIII digestion. The digested cells were further ligated by T4 DNA ligase and reverse crosslinked for DNA purification. A second restriction enzyme NspI digestion was conducted to trim the large chimeric DNA to small fragments of approximately 2 kb, and intramolecular ligated with T4 DNA ligase. After ligation the DNA was purified and used as template to amplify the gene locus of interest by anchor primers.

### Partial digestion and ligation

For the samples implemented by NanoRanger, the digestion reaction was performed with 200ng DNA in a 12.5µl reaction by following the manufacturer’s instructions except that the digestion time was reduced to 5 minutes to achieve stochastic partial digestion that yielded longer restriction DNA fragments. The reaction was then inactivated by incubating for 20 min at 80C, and then placed on ice for 1 min. The ligation reaction of the digested DNA was performed by using T4 DNA ligase following the manufacturer’s instructions. Increasing the reaction volume 10 times is recommended to avoid random ligation events. The DNA was purified by 0.8X AMPure XP beads.

### NanoRanger PCR

For the samples implemented by NanoRanger, the PCR reaction was composed of 10 μl 5X PrimeSTAR GXL Buffer, 4 μl dNTP Mixture, 1 µl inverse primer each (final concentration 10pmol each) (see Supplementary Table 2), 1 μl PrimeSTAR® GXL DNA Polymerase, 10ng purified DNA and sterile purified water to 50 μl. The PCR parameters were set as 95 °C 2 min, repeat 30 times for 98 °C 10 s and then 68 °C 10 min, 68 °C 5 min, 4 °C hold (following the manufacturer’s instructions with the longest recommended extension time).

### Selection of adaptive sampling target regions

16 Bardet Biedl gene loci were selected as gene regions of interest. DNA size distribution was checked using the Agilent Femto Pulse Systems. DNA sample purity was checked using NanoDrop^TM^ 8000 spectrophotometer. For each experiment, The read-N10 of the library was set for the buffer size on either side of the regions of interest. The target bed file was uploaded to MinKNOW software ahead of adaptive sequencing.

### Library preparation and Nanopore sequencing

The native DNA and the products thereafter were quantified using Qubit. Sample products were barcoded and library-prepared using the ONT Ligation Sequencing Kit (SQK-LSK109) and Native Barcoding Expansion 1-12 Kit (EXP-NDB104) following the manufacturer’s instructions (available from https://community.nanoporetech.com/protocols), expect that all elutions were done for 10 min at 37 C. For each library, approximately 50 fmol was loaded onto a Release 9.4.1 flow cell for sequencing on an ONT MinION sequencer. For the samples implemented by adaptive sampling, shearing step was not performed due to the already appropriate fragment size. Approximately 1.2 ug of genomic DNA was used to make sequencing libraries. Sequencing experiments were run for up to 72 h. DNA libraries were loaded onto the same flow cell after washing approximately every 24 h to increase output.

### Sequence analysis

FASTQ files were generated using guppy (v.6.3.4). Sequencing results generated by using NanoRanger approach were analyzed by pyNanoRanger. pyNanoRanger requires input parameters including restriction enzyme sites, inverse primer sequences. pyNanoRanger can operate either in real-time or post-sequencing. It continuously monitors the Nanopore sequencing output folder and processes newly generated fastq files for analysis, sorting them into new folders for each sample in multiplex experiments. If the Nanopore demultiplexing tool Guppy is used, pyNanoRanger performs additional demultiplexing to ensure accurate read classification. pyNanoRanger will select reads that contain the anchor sequences for further analysis. Filtered reads are sorted based on the number of restriction sites they contain and processed accordingly. As sequencing progresses, pyNanoRanger updates sequencing statistics by merging new and existing results. Filtered high-quality long reads were aligned to GRCh38 using minimap2 (v.2.17) with default parameters, enabling users to generate and compare consensus sequences with reference sequences. The complex SVs were identified by searching the variant files generated by Sniffles (v.1.0.12) for SVs that occurred near the suspected mutant locus and by visually checked with IGV (v.2.16.0).

### Genotyping

Uncovered breakpoints were validated by performing PCR and Sanger sequencing. Primer sequence information is attached in Supplementary Table 1. PCR reactions were performed by following the manufacturer’s instructions of the 2× Platinum SuperFi PCR Master Mix (Invitrogen, cat. no. 12358010).

### Large scale singleplex-and multiplex-genotyping

the PCR reaction was composed of 0.2 μl Hot Star Taq polymerase (Qiagen, cat. No. 203205), 2.5 μl of 10X Buffer, 2 μl 2.5 mM dNTP Mixture, 1 µl of each primer (final concentration 10pmol each) (see Supplementary Table 2), 30ng purified DNA and sterile purified water to 25 μl. The PCR parameters were set as 95 °C 10 min, repeat 30 times for 95 °C 30 s, 62 °C 30 s, and then 72 °C 1 min, final extension was set as 72 °C 10 min, and then 4 °C hold.

## Data availability

Raw sequencing data are available in the SRA database (accession ID PRJNA1023960), which is accessible with the following link: https://www.ncbi.nlm.nih.gov/bioproject/PRJNA1023960.

## Code availability

pyNanoRanger is publicly available at https://github.com/YingziZhang-github/pyNanoRanger under the MIT License.

## Acknowledgements

We thank members of Li laboratory for valuable discussion; Jinna Xu, and Doreena Chen for administrative support. We thank Yang Liu of KAUST SRI Center for Uncertainty Quantification in Computational Science and Engineering for developing pyNanoRanger. We thank Changsook Park, Alexander Putra, Angel S. Angelov, and Ptrick P. Driguez of the KAUST Bioscience Core Lab for technical support. Li laboratory was supported by KAUST Office of Sponsored Research (OSR) (BAS/1/1080-01) and KAUST Research Translation Fund Grant (REI/1/4742-01-01).

## Conflict of interest statement

A U.S. Provisional patent application (Application No. 63/588,122) based on methods described in this paper has been filed by King Abdullah University of Science and Technology, in which Y.Z. and M.L. are listed as inventors. The authors declare no other competing interest.

## Author contributions

YZ performed the majority of the molecular biology experiments. CB performed the TLA experiments. YZ, CB, and ML analyzed the data. YZ, FSA, and ML wrote the manuscript. SM, SSN, and MA collected clinical samples. SM, SSN, and MA coordinated the clinical samples and molecular testing. SSN performed Bionano experiments. SM performed the singleplex genotyping in carrier screening. MA performed the multiplex genotyping in carrier screening. ML and FSA conceived the study. FSA and ML supervised the study.

## Notes

### Competing Interest Statement

The authors have declared no competing interest.

### Funding Statement

This study was supported by KAUST Office of Sponsored Research (OSR) (BAS/1/1080-01) and KAUST Research Translation Fund Grant (REI/1/4742-01-01).

### Author Declarations

All human subject samples were collected from King Faisal Specialist Hospital & Research Center (KFSHRC). The research complies with all relevant ethical regulations at KFSHRC. The study of human samples collection was approved by the Institutional Review Board (IRB) of KFSHRC and KAUST Institutional Biosafety and Bioethics Committee (IBEC).

## References

1. RARE Facts – Global Genes [Internet]. *Available from:* https://globalgenes.org/rare-facts/.

2 Graessner, H., Zurek, B., Hoischen, A. & Beltran, S. Solving the unsolved rare diseases in Europe. European Journal of Human Genetics 29, 1319–1320, doi:10.1038/s41431-021-00924-8 (2021).

3 Shashi, V. et al. The utility of the traditional medical genetics diagnostic evaluation in the context of next-generation sequencing for undiagnosed genetic disorders. Genetics in Medicine 16, 176–182, doi:10.1038/gim.2013.99 (2014).

4 Molster, C. et al. Survey of healthcare experiences of Australian adults living with rare diseases. Orphanet J Rare Dis 11, 30, doi:10.1186/s13023-016-0409-z (2016).

5 Monies, D. et al. Lessons Learned from Large-Scale, First-Tier Clinical Exome Sequencing in a Highly Consanguineous Population. Am J Hum Genet 104, 1182–1201, doi:10.1016/j.ajhg.2019.04.011 (2019).

6 Kingsmore, S. F. et al. A Randomized, Controlled Trial of the Analytic and Diagnostic Performance of Singleton and Trio, Rapid Genome and Exome Sequencing in Ill Infants. The American Journal of Human Genetics 105, 719–733, 10.1016/j.ajhg.2019.08.009 (2019).

7 Lowther, C. et al. Systematic evaluation of genome sequencing for the diagnostic assessment of autism spectrum disorder and fetal structural anomalies. Am J Hum Genet 110, 1454–1469, doi:10.1016/j.ajhg.2023.07.010 (2023).

8 AlAbdi, L. et al. Diagnostic implications of pitfalls in causal variant identification based on 4577 molecularly characterized families. Nature Communications 14, 5269, doi:10.1038/s41467-023-40909-3 (2023).

9 Shamseldin, H. E. et al. Increasing the sensitivity of clinical exome sequencing through improved filtration strategy. Genet Med 19, 593–598, doi:10.1038/gim.2016.155 (2017).

10 Lupski, J. R. Genomic disorders: structural features of the genome can lead to DNA rearrangements and human disease traits. Trends Genet 14, 417–422, doi:10.1016/s0168-9525(98)01555-8 (1998).

11 Miller, D. E. et al. Targeted long-read sequencing identifies missing disease-causing variation. Am J Hum Genet 108, 1436–1449, doi:10.1016/j.ajhg.2021.06.006 (2021).

12 Miller, D. E. et al. Targeted long-read sequencing identifies missing pathogenic variants in unsolved Werner syndrome cases. J Med Genet 59, 1087–1094, doi:10.1136/jmedgenet-2022-108485 (2022).

13 Shickh, S., Mighton, C., Uleryk, E., Pechlivanoglou, P. & Bombard, Y. The clinical utility of exome and genome sequencing across clinical indications: a systematic review. Human Genetics 140, 1403–1416, doi:10.1007/s00439-021-02331-x (2021).

14. 14 100,000 Genomes Pilot on Rare-Disease Diagnosis in Health Care — Preliminary Report. New England Journal of Medicine 385, 1868–1880, doi:10.1056/NEJMoa2035790 (2021).

15. Hottentot, Q. P., van Min, M., Splinter, E. & White, S. J. Targeted Locus Amplification and Next-Generation Sequencing. Methods Mol Biol 1492, 185–196, doi:10.1007/978-1-4939-6442-0_13 (2017).

16 Payne, A. et al. Readfish enables targeted nanopore sequencing of gigabase-sized genomes. Nature Biotechnology 39, 442–450, doi:10.1038/s41587-020-00746-x (2021).

17 Dekker, J., Rippe, K., Dekker, M. & Kleckner, N. Capturing chromosome conformation. Science 295, 1306–1311 (2002).

18 Zipfel, P. F. et al. Deletion of complement factor H-related genes CFHR1 and CFHR3 is associated with atypical hemolytic uremic syndrome. PLoS Genet 3, e41, doi:10.1371/journal.pgen.0030041 (2007).

19 Martin, S. et al. Nanopore adaptive sampling: a tool for enrichment of low abundance species in metagenomic samples. Genome Biol 23, 11, doi:10.1186/s13059-021-02582-x (2022).

20 Vogt, J. et al. SVA retrotransposon insertion-associated deletion represents a novel mutational mechanism underlying large genomic copy number changes with non-recurrent breakpoints. Genome Biol 15, R80, doi:10.1186/gb-2014-15-6-r80 (2014).

21 Ottaviani, D., LeCain, M. & Sheer, D. The role of microhomology in genomic structural variation. Trends in Genetics 30, 85–94, 10.1016/j.tig.2014.01.001 (2014).

22 Harel, T. & Lupski, J. R. Genomic disorders 20 years on-mechanisms for clinical manifestations. Clin Genet 93, 439–449, doi:10.1111/cge.13146 (2018).

23 Marwaha, S., Knowles, J. W. & Ashley, E. A. A guide for the diagnosis of rare and undiagnosed disease: beyond the exome. Genome Medicine 14, 23, doi:10.1186/s13073-022-01026-w (2022).

24 Zanello, G., Chan, C. H. & Pearce, D. A. Recommendations from the IRDiRC Working Group on methodologies to assess the impact of diagnoses and therapies on rare disease patients. Orphanet J Rare Dis 17, 181, doi:10.1186/s13023-022-02337-2 (2022).

