## Supplementary figures and images for "NanoRanger enables rapid single base-pair resolution of genomic disorders"

### Fig. S1

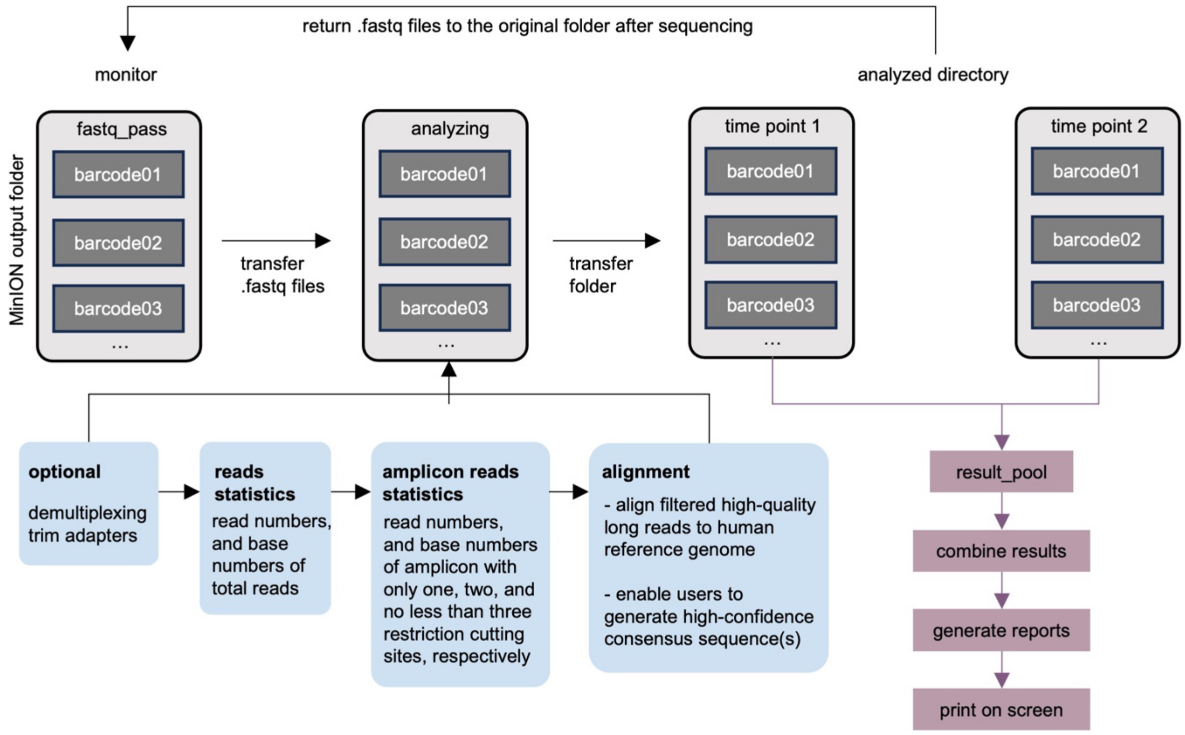


Fig. S1 | Pipeline of pyNanoRanger analysis.
