## Supplementary material for "NanoRanger enables rapid single base-pair resolution of genomic disorders": Fig. S2

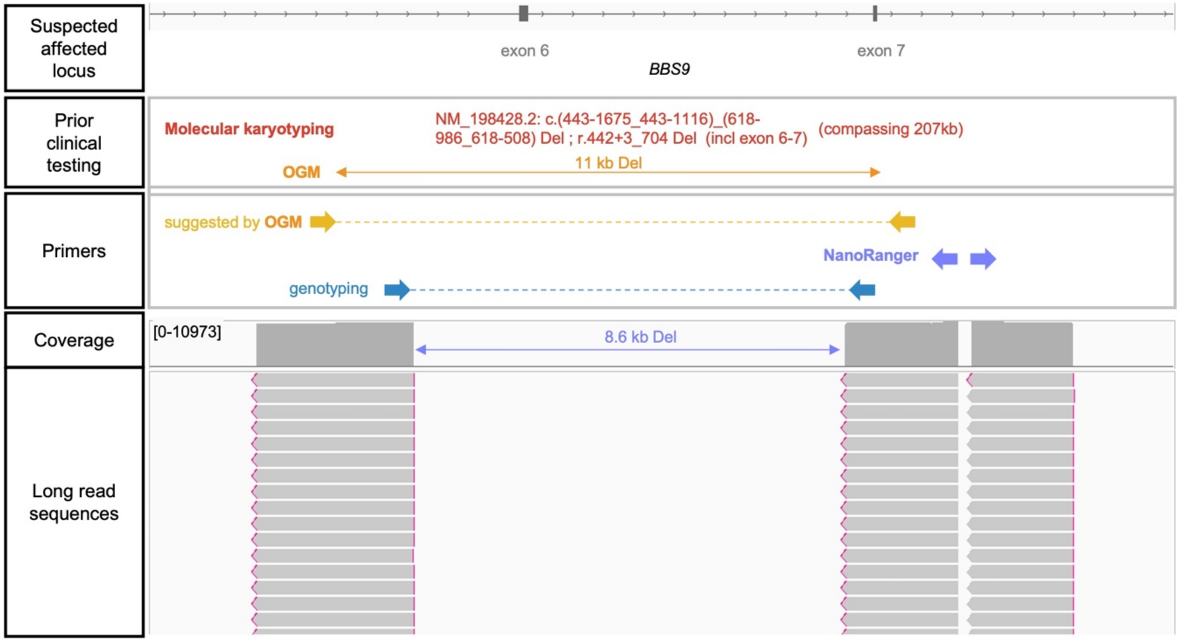


Fig. S2 | NanoRanger enables precise characterization of disease-causing breakpoints in a retinitis pigmentosa case (09DG00509).
