## Supplementary material for "NanoRanger enables rapid single base-pair resolution of genomic disorders": Fig. S3

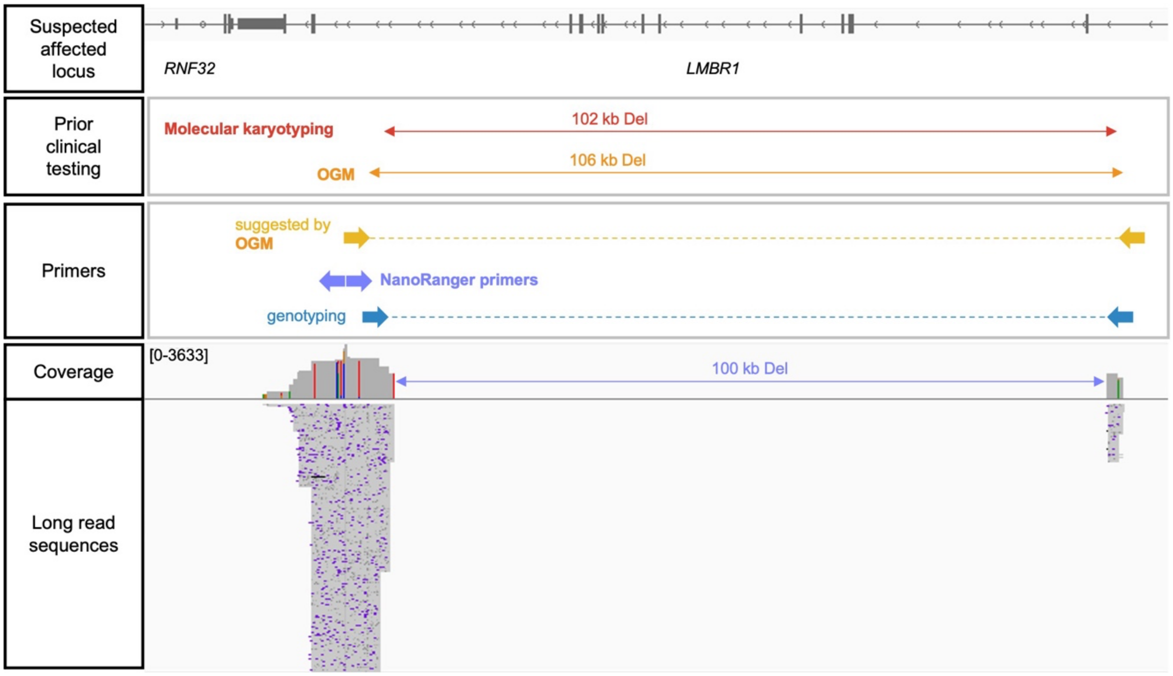


Fig. S3 | NanoRanger enables precise characterization of a female case labeled 14DG0861, characterized by severe developmental defects in both upper and lower limbs.
