## Supplementary material for "NanoRanger enables rapid single base-pair resolution of genomic disorders": Fig. S4

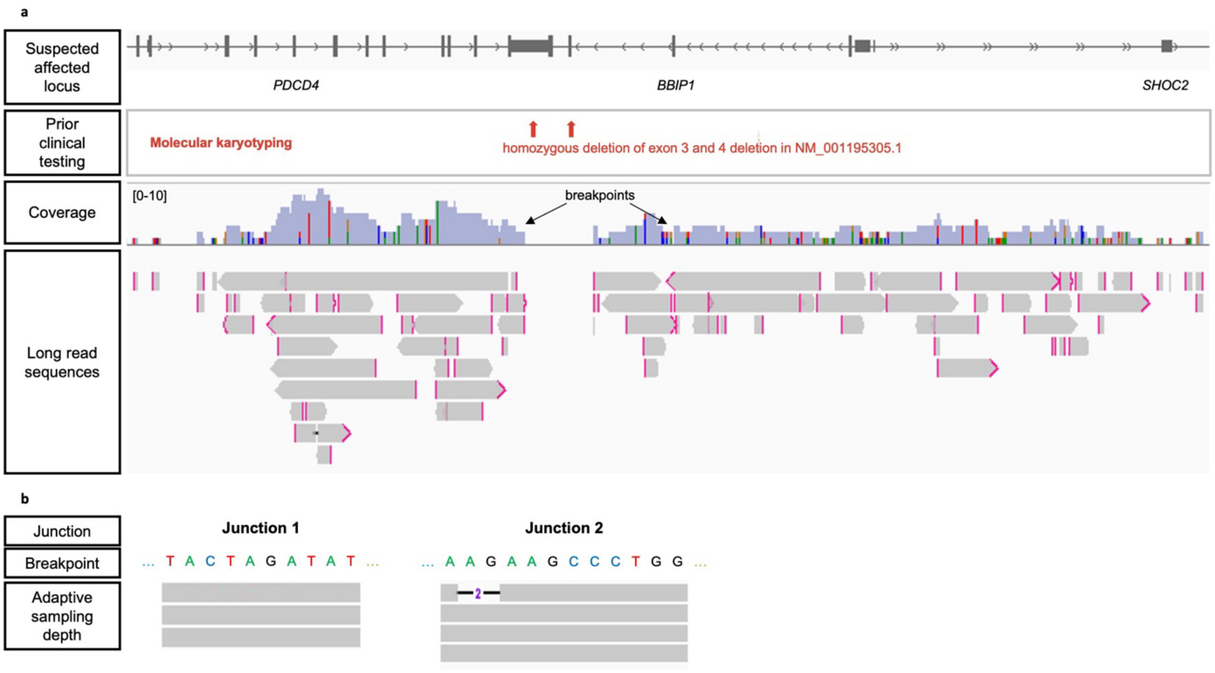


Fig. S4 | Adaptive sampling was applied to 10DG0002 case. **a,** adaptive sampling reads showed the correct breakpoints as identified by NanoRanger. **b,** The two junctions were covered by only three and four reads, respectively, making it difficult to confidently call the true sequence of the SV solely from adaptive sampling.
