## Supplementary material for "NanoRanger enables rapid single base-pair resolution of genomic disorders": Fig. S5

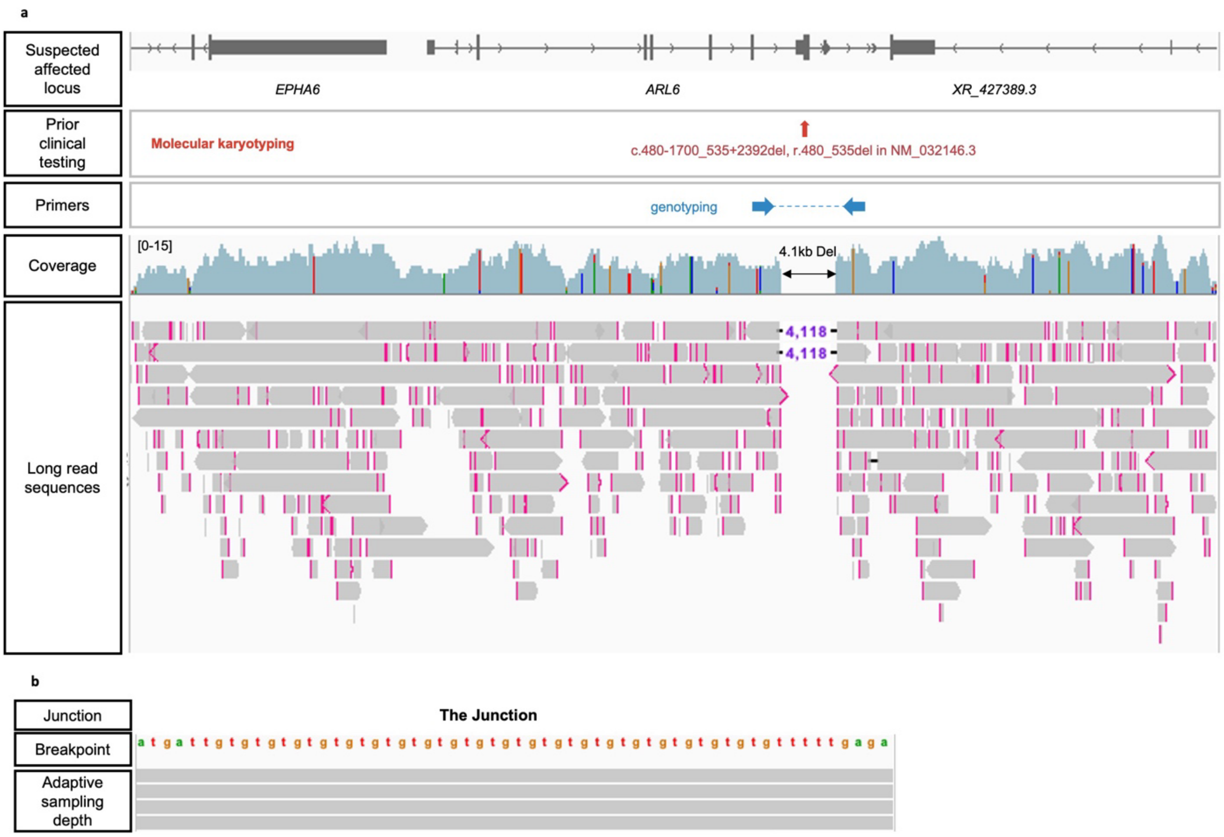


Fig. S5 | Adaptive sampling was applied to 09DG0002 sample. **a,** Adaptive sequencing revealed a large deletion in *ARL6*. **b,** The deletion was covered by four reads.
