## Supplementary material for "NanoRanger enables rapid single base-pair resolution of genomic disorders": Fig. S6

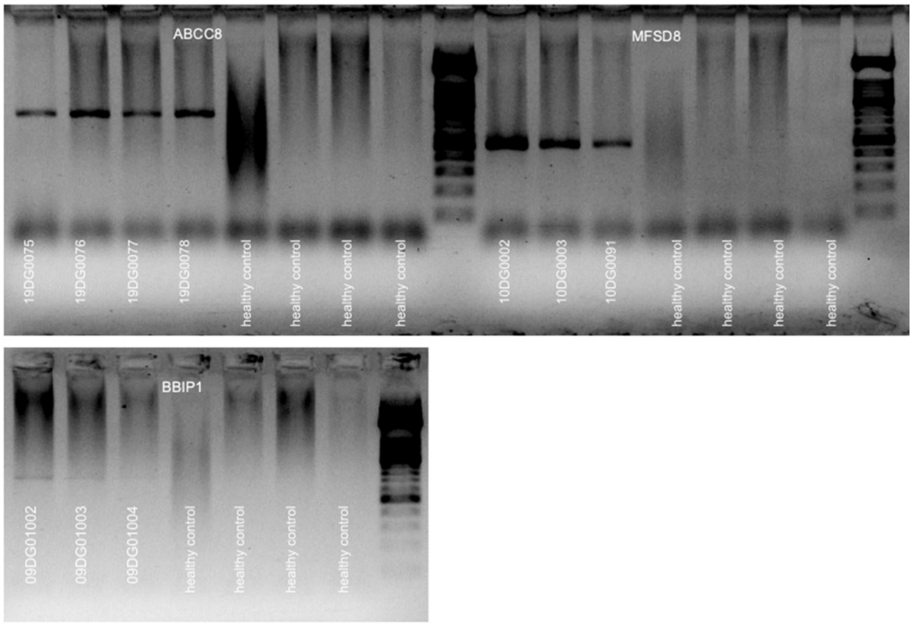


Fig. S6 | The gel electrophoresis of the multiplex genotyping successfully identified the carrier siblings.
