## Supplementary Table 1 for "NanoRanger enables rapid single base-pair resolution of genomic disorders"

Supplementary Table 1 Primer sequence used in this study.

| Oligo | Sequence |
|---|---|
| 17DG0332 LR-PCR forward primer | AGCATTATAAGAGCCGATGGAG |
| 17DG0332 LR-PCR reverse primer | CACGAGCAACCAGCATGTAG |
| 17DG0332 Genotyping forward primer | CGCCTCCAACTCTCAAAGCAA |
| 17DG0332 Genotyping reverse primer | TGCCTCTAACCTCAACTTATT |
| 19DG0075 TLA inverse primer 1 | AGCTGGAAAGAGCCGCGACC |
| 19DG0075 TLA inverse primer 2 | GTGCAACGACGCAGCTGGACCT |
| 19DG0075 TLA inverse primer 3 | GTGGTTCTCGCTGCCGCAGA |
| 19DG0075 TLA inverse primer 4 | TGCCGCACGTCTTCCTACTCT |
| 19DG0075 TLA inverse primer 5 | CCCGCTTCAGGACGATCACCA |
| 19DG0075 TLA inverse primer 6 | AGAAGCTGTGTGAGCAAAAGCCT |
| 19DG0075 TLA inverse primer 7 | ACGATCACCAGGTCTGCACTCA |
| 19DG0075 TLA inverse primer 8 | ATCCCACATTCGGACCCTGC |
| 19DG0075 TLA inverse primer 9 | TCGATGGCCACCAGGACCATG |
| 19DG0075 TLA inverse primer 10 | CAAGTGGACCGACAGCGCCCTGA |
| 19DG0075 TLA inverse primer 11 | TCCAGAGCTGAGAAGGGGTCATC |
| 19DG0075 TLA inverse primer 12 | TAGTGACCCACAAGCTACAGTACC |
| 19DG0075 TLA inverse primer 13 | AGCGGCTCAGGCACTCCAG |
| 19DG0075 TLA inverse primer 14 | TACTTCCGGGTGGCGTCCAG |
| 19DG0075 TLA inverse primer 15 | CTGGCTGAAATTCTCCCCGCCTT |
| 19DG0075 TLA inverse primer 16 | CCTTCGTGAGGAAGACCAGCATCT |
| 19DG0075 TLA inverse primer 17 | ATCCAAGTCGGTCGCTGTCTC |

| | |
|---|---|
| 19DG0075 TLA inverse primer 18 | ATCAGGTACTGCGTCCTGG |
| 19DG0075 Genotyping forward primer | ACCAGCCTGAAGCTCAAAGAGGGC |
| 19DG0075 Genotyping reverse primer | AGGGTGGATGCTCACGGCTCCT |
| 10DG0002 NanoRanger inverse primer 1 | ACAGCCTATGCCCCATTTTGG |
| 10DG0002 NanoRanger inverse primer 2 | CGAAGGAGATGGAGGTCGTC |
| 10DG0002 Genotyping 1 forward primer | CATGATGGTTTATCCACAGGTCCA |
| 10DG0002 Genotyping 1 reverse primer | GCTGCTGTTGAGATACTGTGC |
| 10DG0002 Genotyping 2 forward primer | AGTTCAGAGTAGCTGGAGTTGC |
| 10DG0002 Genotyping 2 reverse primer | TCCTCCCTTTCAGTGGCATATCA |
| 14DG0861 NanoRanger inverse primer 1 | GCTTGGAGCATAAGGATGACACA |
| 14DG0861 NanoRanger inverse primer 2 | GAACTTAGGGAGATGGCTGGA |
| 14DG0861 Genotyping forward primer | GTCACCTCTTGAATGCTTTGCT |
| 14DG0861 Genotyping reverse primer | GACCACAGGCAGAATGGGCTTA |
| 07-00796/07-00462 NanoRanger inverse primer 1 | GCACTGCCTTTGCTGTTCATT |
| 07-00796/07-00462 NanoRanger inverse primer 2 | GGTCACTCACTCTCAAGCCAG |
| 07-00796/07-00462 Genotyping forward primer | GTCCTGCTCCAGGAATTAAACGT |
| 07-00796/07-00462 Genotyping reverse primer | AGCCGTTGCCATCATTCTGA |
| 10DG1265 NanoRanger 1 inverse primer 1 | ATTTACACTTGTGCCCCCGT |
| 10DG1265 NanoRanger 1 inverse primer 2 | GGCTCAGCGATGTCCCTAAA |
| 10DG1265 NanoRanger 2 inverse primer 1 | TGGAGACCGACTAATTTTCTGT |
| 10DG1265 NanoRanger 2 inverse primer 2 | ACTGGCAGAGGAAATAGTCCC |
| 10DG1265 Genotyping forward primer | CTGTAACAGAGCCCAAGGCA |

| | |
|---|---|
| 10DG1265 Genotyping reverse primer | AATGGGCTGCTTCCCTTACC |
| 09DG00509 NanoRanger inverse primer 1 | GCAGGAATGTGATACCATGGAGC |
| 09DG00509 NanoRanger inverse primer 2 | ACACCACTATTGAGGAGGTCAAAGG |
| 09DG00509 Genotyping forward primer | CCTGTTGGCGATTTGTATGTCTTAT |
| 09DG00509 Genotyping reverse primer | GCAAAGCAGTCAGATGGAGTAG |
| 15DG1177 NanoRanger inverse primer 1 | GGCATGTCTGTGGTAATGAGAG |
| 15DG1177 NanoRanger inverse primer 2 | GCAACCTCAGAAGGAGGCCC |
| 15DG1177/15DG11778 Genotyping forward primer | ACGGACAGCAAAGTTTGGGA |
| 15DG1177/15DG11778 Genotyping reverse primer | TCTCAGGCACCATTGTGGTC |
| 12DG0797 NanoRanger 1 inverse primer 1 | TTACAGGCCAGCACGATTCAT |
| 12DG0797 NanoRanger 1 inverse primer 2 | CCAAGCCCAGAAGCAGGTAG |
| 12DG0797 NanoRanger 2 inverse primer 1 | GCCACTCCCAAATCAATAGCA |
| 12DG0797 NanoRanger 2 inverse primer 2 | GGTGATGTAACTGCCATACAAT |
| 12DG0797 Genotyping forward primer | GCCAATTTGTGGTAAAGTCAGGAA |
| 12DG0797 Genotyping reverse primer | TTTTCGGAAAGGAGCTCACA |
| 20DG1339 NanoRanger inverse primer 1 | ACAATGAGCCTCAGAAGCTGT |
| 20DG1339 NanoRanger inverse primer 2 | GTCGGGGTAAAAGTTAGGGTTT |
| 09DG01002 Genotyping forward primer | GTCTCCTGCCTTGGGTACAA |
| 09DG01002 Genotyping reverse primer | ACCCTTCACAGCTACATTCACAT |
