## Supplementary Table 2 for "NanoRanger enables rapid single base-pair resolution of genomic disorders"

Supplementary Table 2 Adaptive sampling data for 293T cell sample control, per sequencing targets, coverage, and average read length.

| Target region | Gene region of interest | Size of gene region of interest | Read counts of interested gene region | Average coverage of interested gene region (x) | Average length of reads in interested gene region (bp) |
|---|---|---|---|---|---|
| *BBIP1* | chr10:110898730-110919201 | 20471 | 37 | 17.3 | 9,578 |
| *BBS1* | chr11:66510606-66533613 | 23007 | 58 | 19.2 | 7,595 |
| *ARL6/BBS3* | chr3:97764521-97801229 | 36708 | 92 | 21.0 | 8,392 |
| *BBS2* | chr16:56582667-56465640 | 117027 | 187 | 11.4 | 7,103 |
| *BBS4* | chr15:72686207-72738473 | 52266 | 119 | 20.3 | 8,917 |
| *BBS5* | chr2:169479480-169506655 | 27175 | 60 | 19.2 | 8,712 |
| *BBS7* | chr4:121824329-121870474 | 46145 | 66 | 12.4 | 8,654 |
| *TTC8/BBS8* | chr14:88824153-88881079 | 56926 | 89 | 10.6 | 6,775 |
| *BBS10* | chr12:76344474-76348415 | 3941 | 13 | 44.9 | 13,614 |
| *TRIM32/BBS11* | chr9:116687305-116701300 | 13995 | 35 | 22.6 | 9,052 |
| *CCDC28B* | chr1:32196011-32205453 | 9442 | 38 | 36.8 | 9,146 |
| *CEP290* | chr12:88049016-88142088 | 93072 | 141 | 11.3 | 7,473 |
| *TMEM67* | chr8:93754844-93832653 | 77809 | 147 | 13.6 | 7,217 |
| *MKS1* | chr17:58205441-58219605 | 14164 | 25 | 19.0 | 10,754 |
| *MKKS* | chr20:10401009-10434222 | 33213 | 59 | 17.0 | 9,559 |

| | | | | | |
|---|---|---|---|---|---|
| *BBS12* | chr4:122700442-122744942 | 44500 | 75 | 10.0 | 5,933 |
