## Supplementary Table 3 for "NanoRanger enables rapid single base-pair resolution of genomic disorders"

Supplementary Table 3 Adaptive sampling data for 10DG0002, per sequencing targets, coverage, and average read length.

| Target region | Gene region of interest | Size of gene region of interest | Read counts of interested gene region | Average coverage of interested gene region (x) | Average length of reads in interested gene region (bp) |
|---|---|---|---|---|---|
| *BBIP1* | chr10:110898730-110919201 | 20471 | 19 | 2.3 | 2426 |
| *BBS1* | chr11:66510606-66533613 | 23007 | 43 | 3.8 | 2029 |
| *ARL6/BBS3* | chr3:97764521-97801229 | 36708 | 65 | 3.7 | 2063 |
| *BBS2* | chr16:56465640-56582667 | 117027 | 202 | 4.2 | 2441 |
| *BBS4* | chr15:72686207-72738473 | 52266 | 99 | 5.5 | 2882 |
| *BBS5* | chr2:169479480-169506655 | 27175 | 58 | 6.2 | 2913 |
| *BBS7* | chr4:121824329-121870474 | 46145 | 73 | 4.6 | 2888 |
| *TTC8/BBS8* | chr14:88824153-88881079 | 56926 | 107 | 4.5 | 2409 |
| *BBS10* | chr12:76344474-76348415 | 3941 | 8 | 3.8 | 1854 |
| *TRIM32/BBS11* | chr9:116687305-116701300 | 13995 | 41 | 8.4 | 2854 |
| *CCDC28B* | chr1:32196011-32205453 | 9442 | 24 | 7.4 | 2904 |
| *CEP290* | chr12:88049016-88142088 | 93072 | 170 | 4.0 | 2175 |
| *TMEM67* | chr8:93754844-93832653 | 77809 | 152 | 4.4 | 2252 |
| *MKS1* | chr17:58205441-58219605 | 14164 | 23 | 4.4 | 2725 |
| *MKKS* | chr20:10401009-10434222 | 33213 | 61 | 4.7 | 2567 |

| | | | | | |
|---|---|---|---|---|---|
| *BBS12* | chr4:122700442-122744942 | 44500 | 88 | 4.6 | 2300 |
