## Supplementary Table 4 for "NanoRanger enables rapid single base-pair resolution of genomic disorders"

Supplementary Table 4 Adaptive sampling data for 09DG0002, per sequencing targets, coverage, and average read length.

| **Target region** | **Gene region of interest** | **Size of gene region of interest** | **Read counts of interested gene region** | **Average coverage of interested gene region (x)** | **Average length of reads in interested gene region (bp)** |
|---|---|---|---|---|---|
| *BBIP1* | chr10:110898730-110919201 | 20471 | 106 | 7.9 | 1532 |
| *BBS1* | chr11:66510606-66533613 | 23007 | 132 | 10.5 | 1821 |
| *ARL6/BBS3* | chr3:97764521-97801229 | 36708 | 185 | 8.2 | 1627 |
| *BBS2* | chr16:56465640-56582667 | 117027 | 595 | 7.4 | 1450 |
| *BBS4* | chr15:72686207-72738473 | 52266 | 266 | 7.5 | 1482 |
| *BBS5* | chr2:169479480-169506655 | 27175 | 140 | 7.6 | 1469 |
| *BBS7* | chr4:121824329-121870474 | 46145 | 218 | 7.3 | 1540 |
| *TTC8/BBS8* | chr14:88824153-88881079 | 56926 | 292 | 8.2 | 1603 |
| *BBS10* | chr12:76344474-76348415 | 3941 | 27 | 10.1 | 1475 |
| *TRIM32/BBS11* | chr9:116687305-116701300 | 13995 | 72 | 8.5 | 1656 |
| *CCDC28B* | chr1:32196011-32205453 | 9442 | 60 | 11.1 | 1738 |
| *CEP290* | chr12:88049016-88142088 | 93072 | 451 | 7.3 | 1496 |
| *TMEM67* | chr8:93754844-93832653 | 77809 | 403 | 7.9 | 1519 |
| *MKS1* | chr17:58205441-58219605 | 14164 | 72 | 10.4 | 2035 |

| | | | | | |
|---|---|---|---|---|---|
| *MKKS* | chr20:10401009-10434222 | 33213 | 193 | 8.7 | 1494 |
| *BBS12* | chr4:122700442-122744942 | 44500 | 251 | 9.4 | 1664 |
