## Supplementary Table 5 for "NanoRanger enables rapid single base-pair resolution of genomic disorders"

Supplementary Table 5 Sequencing data for multiplex NanoRanger test.

| | **0.2 uM** | **0.3 uM** | **0.4 uM** | **0.5 uM** |
|---|---|---|---|---|
| Total reads | 236056 | 231739 | 323003 | 305581 |
| Target 1 *(BBIP1)* | 7448 (3.16%) | 1741 (0.75%) | 2215 (0.69%) | 1483 (0.49%) |
| Target 2 *(ABCC8*) | 3379 (1.43%) | 1480 (0.64%) | 1548 (0.48%) | 1227 (0.40%) |
